# Knowledge and attitude of Iranian university students toward genital warts

**DOI:** 10.1101/2021.04.15.21255532

**Authors:** Zeinab Aryanian, Maedeh Mohammadi, Arefeh Babazadeh, Ifa Etesami, Parvaneh Hatami, Mohammad Barary, Soheil Ebrahimpour, Parisa Sabbagh, Azadeh Goodarzi

## Abstract

Human papillomavirus (HPV) is one of the well-known causes of cervical cancer and one of the most prevalent sexually transmitted diseases (STDs). Since cervical cancer is one of the most common causes of morbidity and mortality in women, this study was performed to investigate students’ knowledge and attitudes in Babol University of Medical Sciences, Northern Iran, towards genital warts. This cross-sectional study assessed the perception and attitudes of 385 students at Babol University of Medical Sciences using a preconceived questionnaire. The data collection tool used in this study was a questionnaire that was confirmed to be valid and reliable. The participants’ mean age was 23.70 ± 3.27 years, with a range of 20-50 years. Evaluation of these students’ level of general information about HPV showed that 7 (1.8%) students had inadequate general information, 34 (8.8%) had moderate knowledge, and 344 (89.4%) had good general knowledge in this setting. There was also a significant association between students’ general knowledge of HPV and their field of study. Based on the present study results, the knowledge of the majority of Babol University of Medical Sciences students about genital warts was in a good range, and their knowledge about HPV routes of transmission was of moderate level. Moreover, the majority had an appropriate attitude to interact with people infected with HPV.

## Introduction

Human papillomavirus (HPV) is the standard warts agent, manifested by multiple skin-colored papules in different parts of the body, including the anus, vulva, penis, and pubis. It may occur in all human populations and can sometimes lead to cancer. Its infectivity mechanism involves the cutaneous and mucosal epithelial tissues from different anatomical regions.^1^ To date, more than a hundred different types of HPV have been identified, one-third of which infect the epithelial cells of the reproductive system.^2^ Human genital papillomavirus types are divided into two categories: low-risk types, such as HPV types 6, and 11, which are the causes of benign warts, do not progress to cancer, and usually improve spontaneously. On the other hand, high-risk types, including HPV types 16, 18, 31, 33, and 45, associated with the development of genital cancer, have been engaged in 99% of cervical cancers, among which HPV 16 is the most common type involved.^3–5^

The increasing prevalence of HPV as the most common sexually transmitted disease (STD) in young people has caused considerable interest in human health.^6^ HPV can be transmitted through sexual intercourse. Studies have shown that 30-60% of sexually active adults become infected with HPV at least once during their reproductive age, and this rate is higher in younger women than the older ones.^7–9^ Nearly 80% of HPV infections disappear within 3 to 4 years without intervention. However, primary prevention using vaccination and screening for cervical cancer appears to be beneficial.^8^ The HPV vaccine is safe and reduces cervical cancer progression in 70% of cases, and to have the utmost efficacy, it should be given before first sexual intercourse.

Nevertheless, recommending the vaccine to a single adult girl has posed challenges.^8,9^ On the other hand, the anxiety of being infected and being diagnosed with HPV infection through Papanicolaou (Pap) smear screening test seems to be due to a lack of knowledge about HPV infection.^10^ Therefore, being aware of the relationship between HPV and cervical cancer is the most critical issue for the general population. Surprisingly, even health care personnel appear to have little information about this entity.^8^ This may be a reasonable cause of public ignorance regarding HPV.^11^ Although vaccine production and routine vaccination programs can promote public awareness of this virus and its complications, there are no such programs in Iran.^8^ Therefore, medical students need such information to improve their knowledge and health status and form other social campaigns to educate the public. Unfortunately, studies in developed countries have shown that most adults and young women, even those with high education, have very little information about HPV infection.^8^ Thus, it is necessary to evaluate the student’s attitudes and knowledge regarding HPV to modify and optimize government health education programs.

Hence, due to the increasing prevalence of genital warts in recent years and the lack of studies to examine medical students’ knowledge and attitudes towards genital warts and their prevention methods and routes of transmission, we aimed to conduct this study at Babol University of Medical Sciences.

## Materials and Methods

This cross-sectional study assessed the perception and attitudes of 385 students at Babol University of Medical Sciences using a pre-established questionnaire. Participants were randomly selected among operating room technicians, nursing, midwifery, lab scientists, and medical students. This study’s data collection tool was a questionnaire similar to that used in an earlier study conducted by Ghojazadeh et al., for which validity and reliability were confirmed. Seven academic experts of Tabriz University of Medical Sciences approved the validity of this questionnaire, and its reliability was confirmed via a pilot study in which Cronbach’s alpha was 80%.^12^ The questionnaire was made up of 3 sections and 62 questions. The first part of this questionnaire included demographic information on participants (8 items). In the second section, they were asked about their knowledge about HPV (10 items), the association of HPV with cervical cancer (2 items), its predisposing factors (10 items), and routes of transmission (22 items). Participants’ attitudes towards the person infected with genital warts were then examined in the third section (10 items).

The minimum score was 0, and the maximum was 10. To determine the amount of general information in terms of quality and its easier understanding, the interpretation of obtained scores were as follows: Scores of 0-3 were considered inadequate, 4-7 were considered moderate, and 8-10 were considered Favorable.

Finally, the data were analyzed using the SPSS v. 22 software (IBM, Chicago, IL, USA). Chi-square tests were used, and a p-value less than 0.05 was considered statistically significant.

## Results

In the current study, 385 students participated with a mean age of 23.70 ± 3.27 years, with a range of 20-50 years. The students’ average academic semester was 8.71 ± 3.47, with a range of 1-14. Evaluation of students’ level of general knowledge about HPV showed that 7 (1.8%) students had inadequate general information, while 34 (8.8%) and 344 (89.4%) students had moderate and favorable information in this field, respectively. Evaluation of students’ information regarding the predisposing factors of cervical cancer demonstrated that 4 students (1%) had inadequate general information, 176 (45.7%) had moderate information, and 205 (53.2%) had good information. The students’ knowledge about HPV transmission routes and contributing factors was as follows: 201 (52.2%) had moderate knowledge, and 184 (47.8%) had good knowledge (Table 1).

**Table 1.**
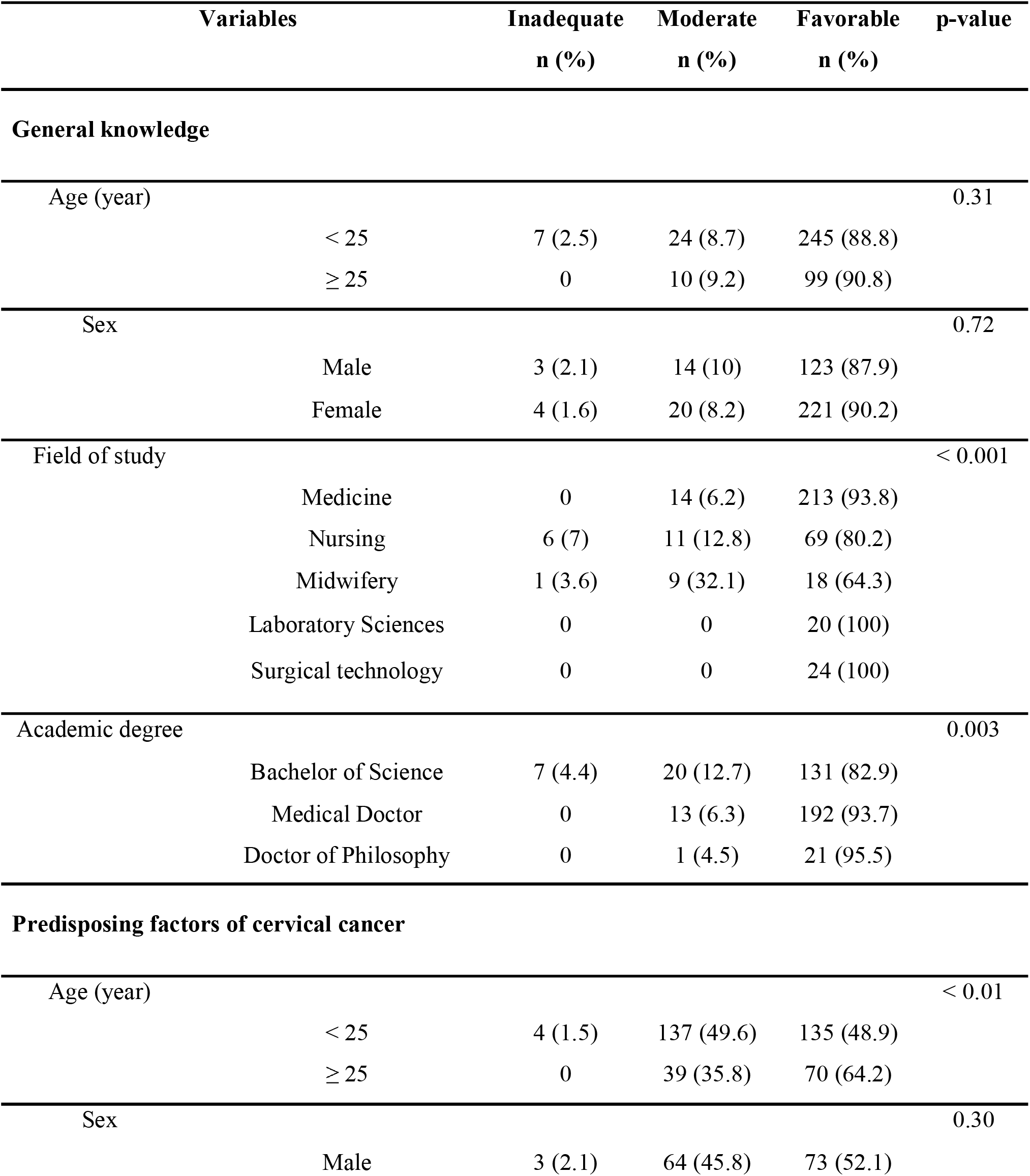
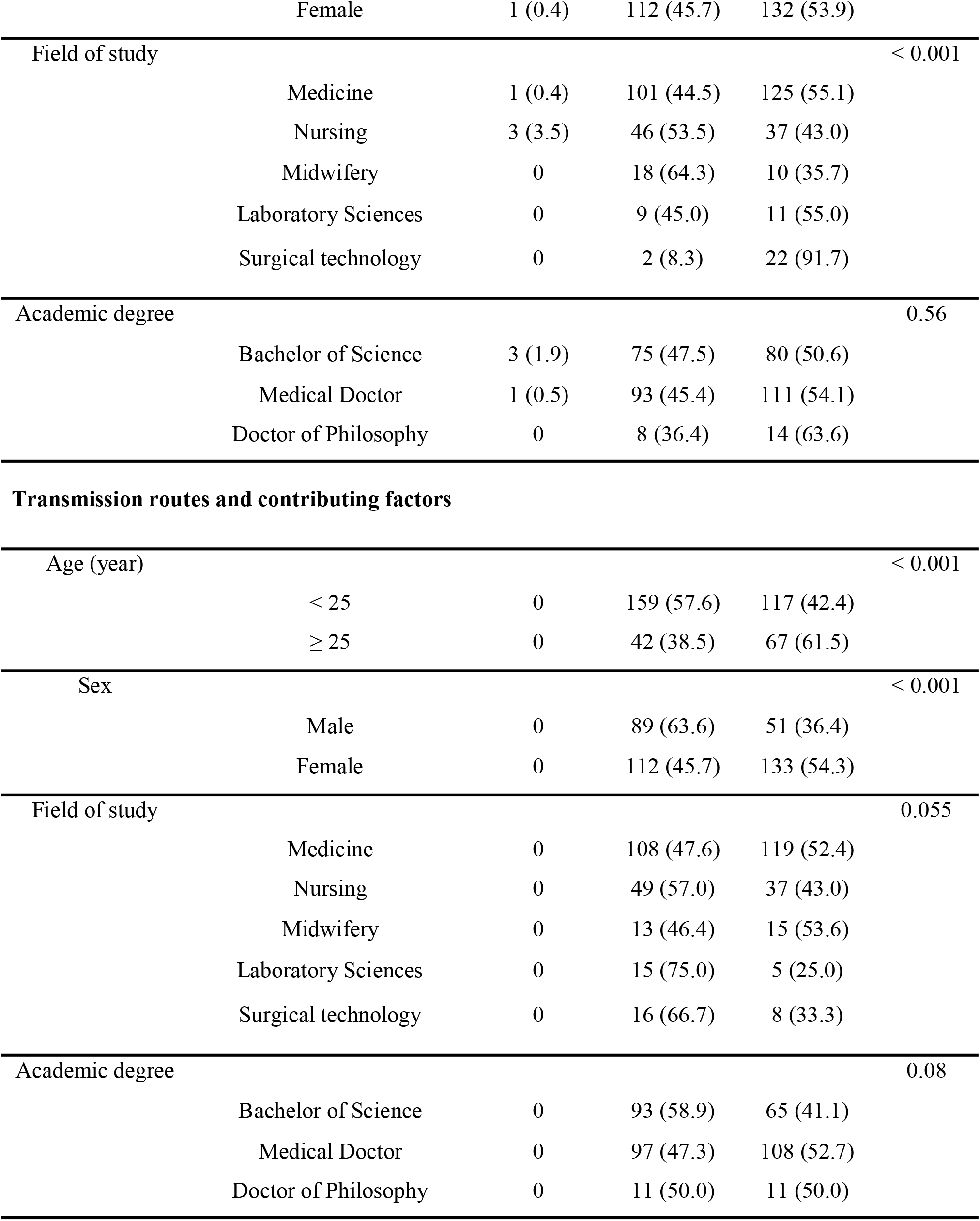
Comparison of students’ general knowledge, predisposing factors of cervical cancer, and transmission routes and contributing factors stratified by their demographic characteristics

According to Table 1, the extent of general information about HPV infection among students is linked to their field of study and education level (p < 0.001 and p = 0.003, respectively). However, no significant correlation between the students’ knowledge and age or sex was observed. Moreover, the extent of students’ information regarding the predisposing factors of cervical cancer was significantly correlated with age and field of study (p < 0.01 and p < 0.001, respectively). Furthermore, regarding the amount of students’ knowledge about the HPV transmission routes and contributing factors, they were significantly correlated with students’ age and sex (p < 0.001 and p < 0.001, respectively).

The attitude of the students about communication with HPV-infected individuals was as follows: 90 participants (23.4%) moderate-quality attitude while 295 (76.6%) had a good attitude (Table 2). According to Table 2, the extent students’ attitude toward HPV-infected patients was significantly correlated with their age, field of study, and academic degree (p < 0.01, p < 0.001, and p = 0.003, respectively).

**Table 2.** Comparison of students’ attitude toward human papillomavirus (HPV)-infected patients stratified by their demographic characteristics

| Variables |  | Poor<br>n (%) | Moderate<br>n (%) | Favorable<br>n (%) | p-value |
| --- | --- | --- | --- | --- | --- |
| Age (year) | < 25 | 0 | 74 (26.8) | 202 (73.2) | < 0.01 |
|  | ≥ 25 | 0 | 16 (14.7) | 93 (85.3) |  |
| Sex | Male | 0 | 31 (22.1) | 109 (77.9) | 0.70 |
|  | Female | 0 | 59 (24.1) | 186 (75.9) |  |
| Field of study | Medicine | 0 | 41 (18.1) | 186 (81.9) | < 0.001 |
|  | Nursing | 0 | 28 (32.6) | 58 (67.4) |  |
|  | Midwifery | 0 | 19 (67.9) | 9 (32.1) |  |
|  | Laboratory Sciences | 0 | 2 (10.0) | 18 (90.0) |  |
|  | Surgical technology | 0 | 0 | 24 (100) |  |
| Academic degree | Bachelor of Science | 0 | 49 (31.0) | 109 (69.0) | 0.003 |
|  | Medical Doctor | 0 | 40 (19.5) | 165 (80.5) |  |
|  | Doctor of Philosophy | 0 | 1 (4.5) | 21 (95.5) |  |

## Discussion

The study’s objective was to investigate Babol University of Medical Sciences students’ knowledge and attitude towards genital warts, their prevention, and transmission pathways. Various analyses of the participants’ knowledge and attitudes toward genital warts showed that almost 90% of the students had good knowledge about sexually transmitted diseases (STDs). Also, 53% of the students had good knowledge about the predisposing factors for cervical cancer. The extent of participant’s education and awareness about HPV pathways and their contributing factors was intermediate among most students. Finally, the vast majority of students had a good attitude about interacting with people infected with HPV.

HPV is one of the most common sexually transmitted infections globally. The primary predisposing factor of cervical cancer in women aged 15 to 44.^13^ The vital issue in the evaluation of students’ knowledge and attitude is the investigation of the student’s level of knowledge about HPV infection, HPV-related risk factors, routes of transmission, and prevention of this infection and consequently the resultant cervical cancer. Various efforts have been undertaken to improve the awareness and knowledge of this group of population. Nevertheless, further studies are needed to raise students’ awareness and knowledge and subsequently improve HPV-infected patients’ attitudes and reduce the high-risk population.

A recent survey on HPV general information revealed that 10.6% of the students did not have good general knowledge about HPV. Since the questions applied in this survey were straightforward and despite this simplicity, some students had little information in this field, preparing educative programs to increase their level of awareness seems reasonably necessary. In another study by Grigore et al. in 2018, it was found that 69.2% of the participating women were found to have information about HPV, but their knowledge in this area was minimal. It has been inferred that the provision of comprehensive education programs could reduce the prevalence of this infection.^14^ Another study conducted by Pourkazemi et al. in 2017 also recognize low awareness among students about HPV infection.^15^ To evaluate the community’s knowledge level about this STD, it might be reasonable first to examine the medical community and medical students since this population will be responsible for the community’s health education. In the present study, the level of awareness, knowledge, and attitude has been above the intermediate level, but in Pourkazemi’s study, this level has been much lower than expected, which could be due to differences in the curricula of the two universities. The exchange of information and teaching courses is done comprehensively and entirely in some universities of medical sciences, leading to the difference in knowledge level in different universities’ students.^15^

Evaluation of the students’ knowledge about HPV, based on age, sex, field of study, and degree of academic education, demonstrated that the students’ general information about HPV had no relationship with their age and sex. However, there was a relationship between their knowledge level and field and degree of study. In general, it appears that midwifery and nursing students had less general information than other medical fields. Nevertheless, Salehifar et al. stated students at the School of Nursing and Midwifery had the highest level of knowledge, which was contrary to the results of this study.^16^ The reason for this disagreement can be attributed to the difference in the academic year and semester of students involved in these studies since with the progression of the academic year, students will learn more about this topic.

Furthermore, undergraduate students had a lower level of knowledge than Ph.D. students and residents of medical specialties. In our study, 46.7% of the students had inadequate information about the contributing factors of cervical cancer, indicating the necessity of organizing a comprehensive program to improve the students’ awareness of cervical cancer risk factors. In 2014, Kwang et al. found that the students’ pre-university knowledge of HPV and cancer-predisposing factors was low. They also noted that interest in vaccination could increase the students’ level of awareness and knowledge.^17^ Besides, the relationship between the field of study and the level of knowledge about cervical cancer’s precipitating factors has been significant. It has been shown that surgical technology students had the most favorable and midwifery students had the lowest level of knowledge. Furthermore, medical and laboratory science students were on the same level, and nursing students in the following ranks.

Evaluation of the relationship between Babol University of Medical Sciences students’ awareness about HPV transmission routes and its contributing factors with essential variables in this study showed that 61.5% of students over 25 years old had good knowledge in this field. In comparison, 57.6% of students under 25 years of age had moderate knowledge, suggesting that aging raises awareness of HPV transmission routes and their contributing factors. Similarly, a 2013 study by Ortashi et al. stated that students are more likely to become aware of the human papillomavirus, its transmission routes, and its predisposing risk for cervical cancer as they age.^18^ Female students also had a higher level of awareness compared to men. The reason may be because women are precisely the population at risk for cervical cancer by HPV. Therefore, knowledge of transmission pathways and risk factors seems more vital to women than to men.

In the present study, over 70% of students had a good attitude towards interacting with people infected with HPV. Furthermore, older students have a better attitude toward this topic. Also, surgical technology, laboratory science, and medical students had the most optimistic interactions with patients. According to the results, midwifery students did not have a good attitude towards people infected with HPV, which was expectable due to the constant interaction of midwives with infected women. Nonetheless, considering that midwifery students are the front line in treating gynecological diseases, it seems very important to improve their attitude.

Interestingly, medical residents had the best attitude toward this issue. This finding was in agreement with Ghojazadeh et al., where residents were more experienced in contacting infected patients than other students.

It is recommended that other studies should be conducted to evaluate students’ knowledge and attitude towards the HPV vaccine. Also, such studies in the general population would be of interest.

## Conclusion

Based on the present study results, the knowledge level of most students of Babol University of Medical Sciences about genital warts was good, but their information about the routes of transmission was intermediate. Furthermore, it is suggested that educational programs should be modified and optimized to improve awareness among teachers, health workers, nurses, and medical staff of non-medical universities.

## Data Availability

The data that support the findings of this study are available from the corresponding author upon reasonable request.

## Acknowledgments

The authors thank the Department of Infectious Diseases of Babol University of Medical Sciences.

